# Proinflammatory Cytokine Levels in Sepsis and in Health and TNFα Association with Sepsis Mortality and patient characteristics: a Systematic Review and Meta-analysis

**DOI:** 10.1101/2021.12.13.21267720

**Authors:** Amal A. Gharamti, Omar Samara, Anthony Monzon, Gabrielle Montalbano, Sias Scherger, Kristen DeSanto, Daniel B. Chastain, Stefan Sillau, Jose G. Montoya, Carlos Franco-Paredes, Andrés F. Henao-Martínez, Leland Shapiro

## Abstract

**Background:** Sepsis is a global health problem associated with significant morbidity and mortality. Detrimental sepsis effects are attributed to a “cytokine storm.” However, anti-cytokine therapies have failed to lower sepsis mortality. We aim to characterize levels of key cytokines in sepsis patients and healthy controls and relate TNFα levels to patient characteristics and outcomes.

**Methods:** We performed a systematic review and meta-analysis. Medline, Embase, Cochrane Library, and Web of Science Core Collection databases were searched from 1985 to May 2020 for studies in English. We included randomized controlled trials (RCTs), controlled trials, cohort studies, case series, and cross-sectional studies that reported mean levels of cytokines in the circulation thought to be relevant for sepsis pathogenesis. We also evaluated concentrations of these cytokines in healthy persons. Quality in Prognosis Studies tool was used to assess the methodological quality of included studies. We extracted summary data from published reports. Data analyses were performed using a random-effects model to estimate pooled odds ratios (OR) with 95% confidence intervals for cytokine levels and mortality. This systematic review is registered in PROSPERO (CRD42020179800).

**Findings:** We identified 3654 records, and 104 studies were included with a total of 3250 participants. The pooled estimated mean TNFα concentration in sepsis patients was 58.4 pg/ml (95% Confidence Interval or CI 39.8-85.8 pg/ml) and 5.5 pg/ml (95% CI 3.8-8.0 pg/ml) in healthy controls. Pooled estimate means for IL-1β and IFNγ in sepsis patients were 21.8 pg/ml and 63.3 pg/ml, respectively. Elevated TNFα concentrations associated with increased 28-day sepsis mortality (p=0.001). In subgroup analyses, TNFα levels did not relate to sepsis source, sepsis severity, or sequential organ failure assessment (SOFA) score.

**Interpretation:** TNFα concentration in sepsis is increased approximately 10-fold compared to healthy persons, and TNFα associated with sepsis mortality but not with sepsis severity. The concept that elevated cytokines cause sepsis should be revisited in the context of these data.

**Funding:** None.

## Introduction

No concept is more commonly invoked as a cause of disease than hyper inflammation. The prototype disease thought to be caused by excessive inflammation is sepsis. In 2016, the Surviving Sepsis Campaign defined sepsis as “life-threatening organ dysfunction due to a dysregulated host response to infection” [1]. Infections that cause sepsis can be severe enough to cause organ malfunction or death. Annual worldwide estimates include 31.5 million sepsis cases, 19.4 million severe sepsis cases, and 5.3 million deaths [2]. An analysis of hospital mortality in United States hospitals showed that one-third to one-half of inpatient deaths were attributable to sepsis [3]. Sepsis also represents the most costly disease treated in US hospitals [4]. At present, no laboratory test defines sepsis, but three principal clinical sepsis definitions that have been created select similar patient populations[5–7]. As the theoretical cause of sepsis, hyper inflammation is initiated and sustained by excess proinflammatory endogenous molecules (especially cytokines), resulting in a cytokine storm [8]. Substantial research and clinical efforts have been expended to block the bioactivity of cytokines to treat sepsis. However, anti-cytokine therapies to lower sepsis mortality have uniformly failed in clinical trials [9–13].

Tumor necrosis factor (TNF)α, interleukin (IL)-1β, and interferon (IFN)γ are considered relevant mediators of the “cytokine storm” in sepsis [14]. Anti-cytokine therapy failure could be due to inadequate blockade of these cytokines or failure of concept where cytokines are not the cause of sepsis. As a starting point, it is crucial to quantify cytokine levels in sepsis, which will help characterize cytokine role in sepsis pathogenesis.

It is striking that despite decades of research, hundreds of clinical trials, and billions of dollars expended to reduce hyper inflammation in sepsis, no systematic review or meta-analysis has provided firm estimates of circulating levels of relevant cytokines in sepsis. Moreover, no analysis has characterized these cytokine levels in the circulation in healthy volunteers. Without these kinds of studies, we cannot understand the quantitative features of cytokine storm in sepsis, and we lack perspective needed to determine if cytokine elevations in sepsis are, in fact, extraordinary. Understanding levels of proinflammatory cytokines in sepsis will permit meaningful comparisons to levels in other diseases. Regarding cytokine levels in sepsis, one goal is to answer the question, “is that a lot?”

This systematic review aims to quantify key cytokine levels in the circulation in sepsis patients and in healthy volunteers and assess associations between these levels and outcomes and clinical characteristics.

## Methods

### Search Strategy and Selection Criteria

We performed a systematic review and meta-analysis of studies reporting key cytokine levels in sepsis patients and healthy controls. The systematic review considered randomized controlled trials (RCTs), controlled trials, cohort studies, case series, and cross-sectional studies that reported relevant cytokine levels in patient cohorts. We excluded case reports and studies reporting cytokine levels in medians. Only studies published in English were included. Selected studies reported cytokine measurements in venous blood, serum, or plasma and expressed levels in mass units per volume. Quantified data depicted only in graphical form were excluded. We included studies using enzyme-linked immunosorbent assay (ELISA) or bead-based immunoassay technologies. We excluded studies using radioimmunoassay (RIA)-based assays or bioassays. ELISA provides accurate measurement of TNFα in serum, and RIA tends to overestimate TNFα amounts [15].

Enzyme-linked immunosorbent assays and bead-based immunoassays have shown comparable results for circulating cytokine concentrations[16]. We verified that studies included defined sepsis according to the diagnostic criteria proposed by the American College of Chest Physicians and the Society of Critical Care Medicine in 1992 as the presence of systemic inflammatory response syndrome and infection [17]. The 1992 definition was used since most studies reporting cytokine levels used the 1992 definition. The detailed methods and Medline search strategy are available (Supplement. eMethods). Additional details in the systematic review protocol were published (PROSPERO CRD42020179800)[18]. This study followed the Preferred Reporting Items for Systematic Reviews and Meta-analyses (PRISMA) reporting guideline to register the protocol, data collection and integrity, assessment of bias, and sensitivity analyses. A systematic search in Medline (via Ovid), Embase (via Elsevier), Cochrane Library (via Wiley, including Cochrane Central Register of Controlled Trials), and Web of Science Core Collection (via Clarivate Analytics, including Science Citation Index Expanded and Social Sciences Citation Index) databases was conducted on 1 May 2020. Studies published since 1985 written in English were included. Search terms included sepsis, septic shock; purpura fulminans; healthy volunteers, and tumor necrosis factor-alpha.

### Data Analysis and Quality Assessment

Study data were collected and managed using REDCap electronic data capture tools hosted at the University of Colorado Anschutz Medical Campus. We extracted summary data from published reports that included type of study, number of participants, study methods, the technology used to measure three proinflammatory cytokines (TNFα, IL-1β, and IFNγ), and outcomes of interest. We also extracted clinical information in the patient cohorts examined. Clinical variables included mean age, gender, race, the proportion of microbial etiology of sepsis (bacterial, viral, fungal), sepsis severity (sepsis, severe sepsis, or septic shock), and proportion of the site of origin of infection (blood, urine, skin, and soft tissue, respiratory, other). Sources of infection were recorded, allowing the source to overlap if multiple sources were present in a single study. Age was recorded as -1 for preterm infants. The magnitude of organ dysfunction in the cohorts was assessed by recording the mean Sequential Organ Failure Assessment (SOFA) score during the first 24 hours after admission. We compared proinflammatory cytokine concentrations to 28-day mortality. Additional cytokine assessments included association with SOFA score. Critical appraisals were conducted using the Quality in Prognosis Studies (QUIPS) tool [19].

### Statistical Analysis

We extracted the mean and standard deviation for each cytokine concentration in studies and calculated the standard error and log-transformed mean values. We then entered the log-transformed mean and standard errors in a random-effect meta-analysis to combine the log mean values, which were back-transformed for interpretation. Between-study heterogeneity was estimated using the I^2^ statistic, where larger I^2^ indicates increased study heterogeneity. Subgroup and meta-regression analyses were conducted to examine factors associated with cytokine levels and which factors contributed to heterogeneity. Assessed factors included age, gender, the severity of sepsis, and type of sepsis. Continuous variables were dichotomized using the mean. When high heterogeneity was obtained (I^2^≥60%), we performed a subgroup analysis or removed selected studies to assess the source of the high heterogeneity. Linearity between TNFα levels and continuous outcome variables was evaluated using a scatter plot. We also conducted a sensitivity analysis to investigate the effect of individual studies on pooled outcomes. The pooled effect was calculated after excluding one study from the analysis and repeating this single-study exclusion for each study. Contour-enhanced funnel plots were constructed to assess publication bias using the egger test to check for a small-study effect. Statistical analyses were performed using Stata software, version 16.0 (StataCorp). Data were analyzed from December 2020 to January 2021.

## Results

### Literature Search for Sepsis Studies

A total of 3654 records were identified through database searches and were screened for eligibility based on titles and abstracts. Four hundred twenty-seven full-text articles were reviewed. We excluded 323 studies (reasons included cytokine levels not reported in a way that permitted numerical extraction, medians were reported as opposed to means, or cytokine measurement assay did not meet inclusion strategy). After exclusions, 104 studies were included in the meta-analysis with a total of 3250 participants. Sixty-nine studies reported sepsis studies with TNFα levels, 24 studies reported IL-1β levels, and 5 studies disclosed IFNγ (Figure 1).

**Figure 1:**
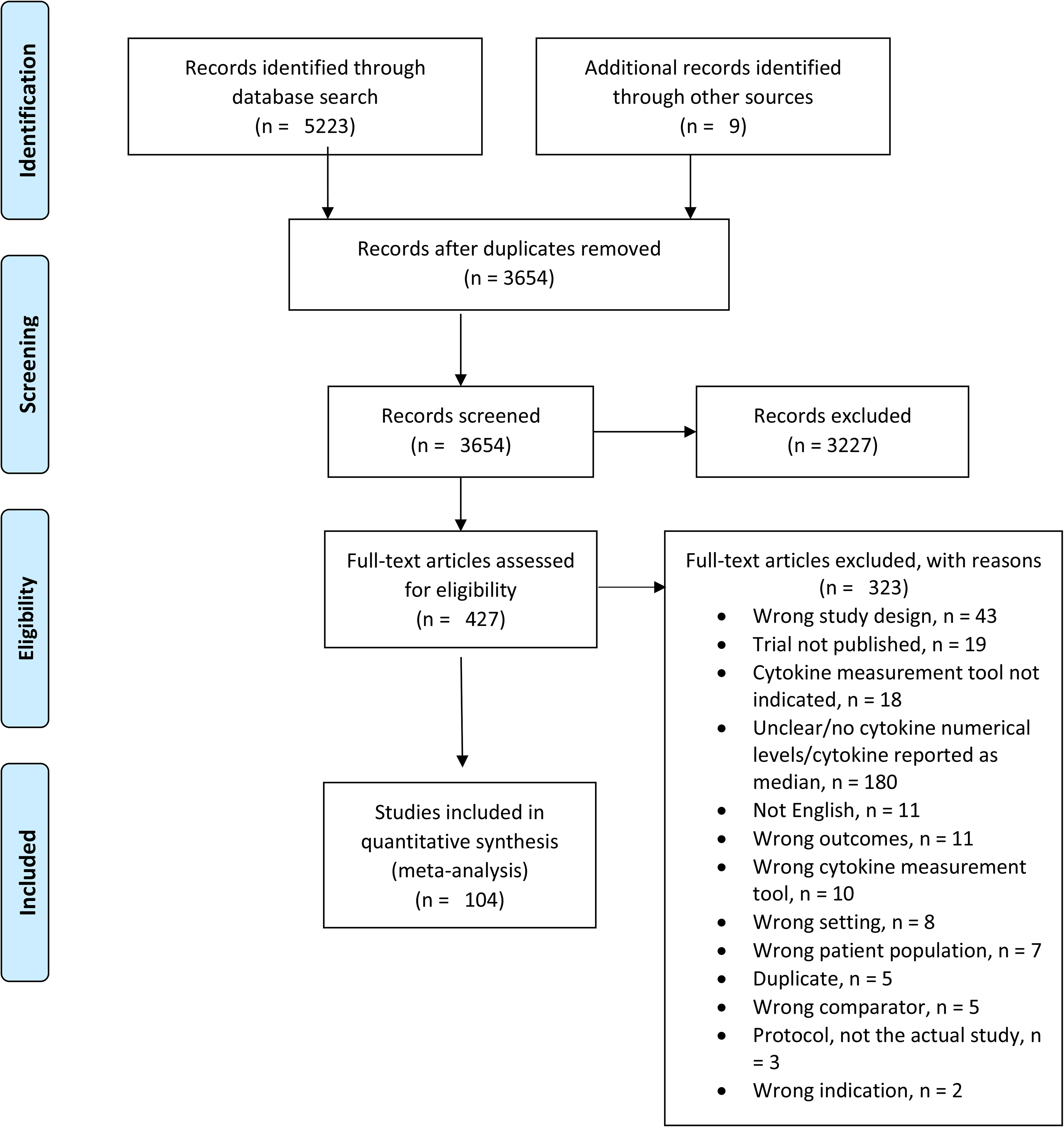
PRISMA Flowchart.

### Literature Search for Healthy Persons Studies

Three hundred ninety-four records were identified through database searches and were screened for eligibility based on titles and abstracts. Seventy full-text articles were reviewed, and 46 were considered for inclusion in the quantitative synthesis. Twelve studies reporting TNFα levels satisfied inclusion criteria.

### TNFα Levels in Sepsis

Sixty-nine studies reporting TNFα levels in sepsis patients were included (Table 1). The mean number of participants in each study was 47.1, ranging between 6 and 220 subjects. Included studies were prospective cohorts (49.3%), randomized clinical trials (37.7%), case-control studies (8.7%), cross-sectional reports (2.9%) and experimental comparative studies (1.5%). Cytokines were quantified using ELISA in 79.7% of studies and using bead-based immunoassay technologies in 20.3% of studies. Most studies were conducted in adults (69.6%), with 30.4% of studies conducted in pediatric patients. The mean age was 42.1 ± 26 years, and ages ranged from preterm infants to 70.3 years. Among adult studies, the mean age was 56.7 ± 9.7 years. The mean percent of female subjects in all included studies was 36.3 ± 11.8. Approximately half of studies reported the microbiologic etiology of sepsis, with the majority originating from bacteria (30.4%), followed by combinations of pathogens (11.6%) and fungal origin (3%). Reported severities of sepsis included sepsis alone (43.5%), severe sepsis (18.8%), or septic shock (37.7%). In studies reporting the source of sepsis, we determined respiratory sources in 33.3%, intra-abdominal infections in 33.3%, and urinary site of origin in 26.1%. The mean SOFA score was 8.5 ± 2.1 in studies that reported SOFA. We could not determine the aggregate frequency of different ethnicities or underlying immunosuppressive conditions due to a lack of reporting. The pooled estimate mean TNFα level in sepsis patients was 58.4 pg/ml (95% CI 39.8-85.8 pg/ml; I^2^ = 99.4% (Figure 2). In studies reporting mortality, the mean mortality at 28 days was 21.9 ± 21.2%, ranging from 5% to 80%.

**Table 1.**
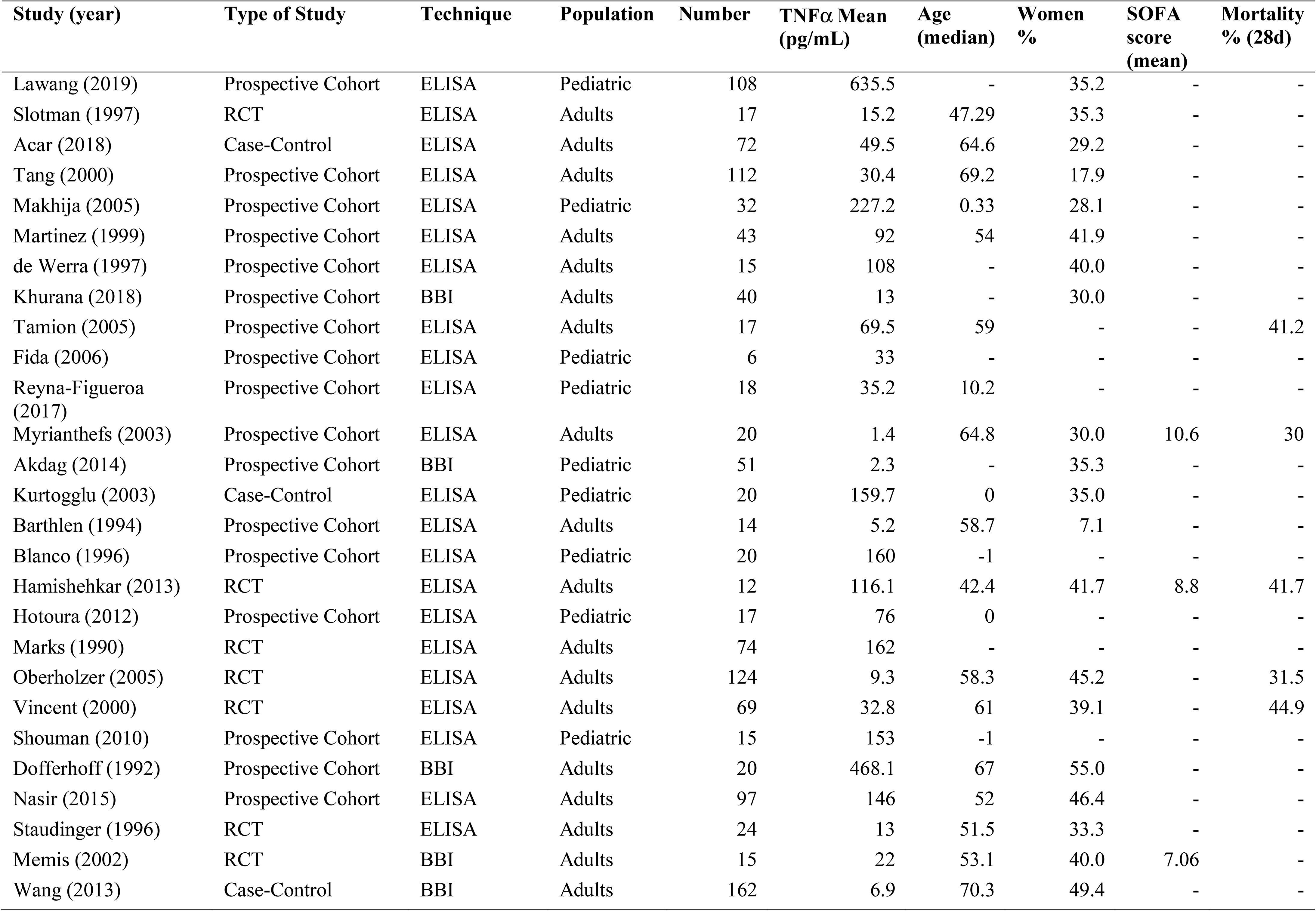
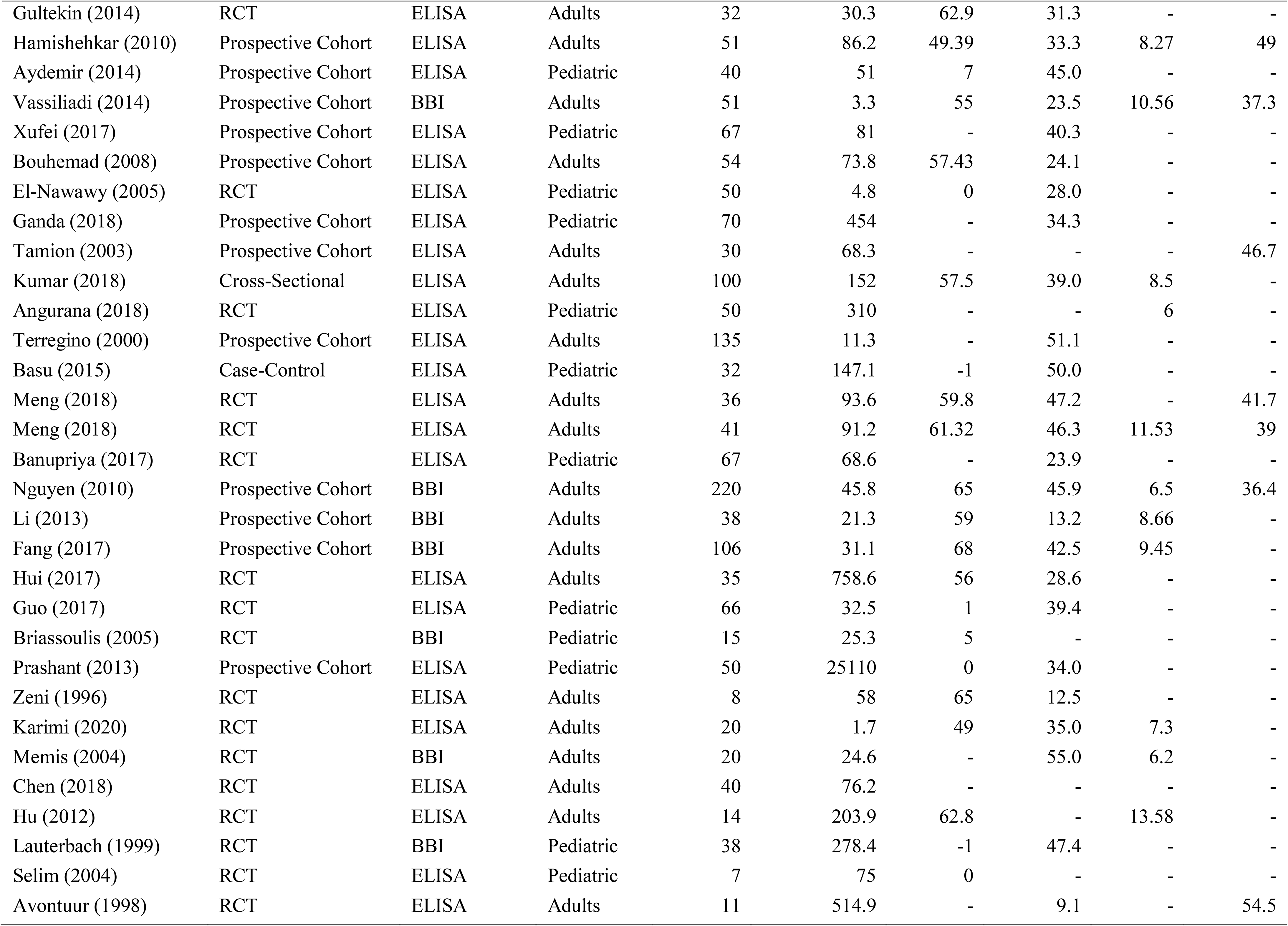
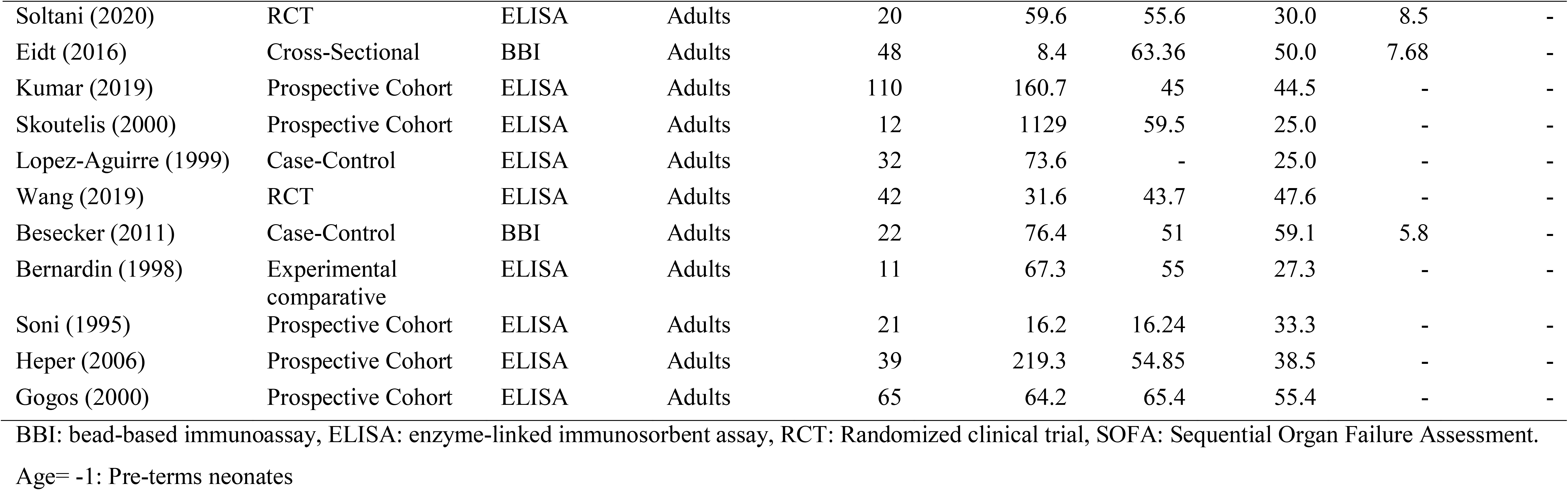
Characteristics of Studies with TNFα levels in sepsis [32–100]

**Figure 2.**
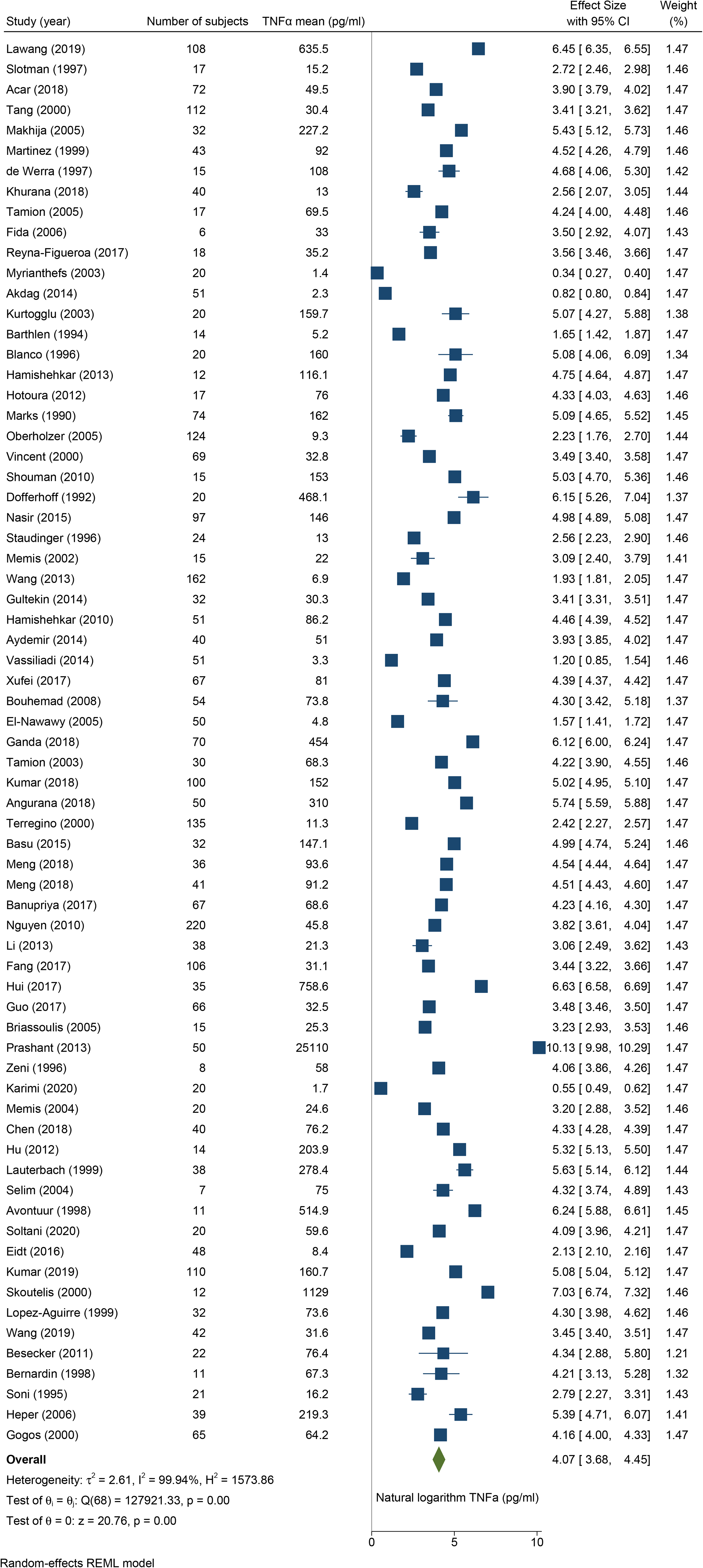
Forest plot for TNFα studies Effect size (pg/ml) is log-transformed.

### TNFα Levels in Sepsis Subgroups

Subgroup analyses indicated no difference in mean TNFα comparing types of studies (66.2 pg/ml for prospective studies, 57.7 pg/ml for case-control studies, 35.8 pg/ml for cross-sectional studies, and 51.5 pg/ml for RCTs; p= 0.929). No significant difference in mean TNFα was noted in adult (45.9 pg/ml) compared to pediatric (101.3 pg/ml) sepsis populations (p=0.086). There were no differences in TNFα levels in sepsis patients with different sites of origin of infection (Figure 3A). Studies measuring TNFα levels using ELISA had significantly higher mean TNFα compared to mean TNFα in studies using bead-based immunoassays (73.5 pg/ml vs. 23.3 pg/ml, respectively; p= 0.011) (Figure 3B).

**Figure 3:**
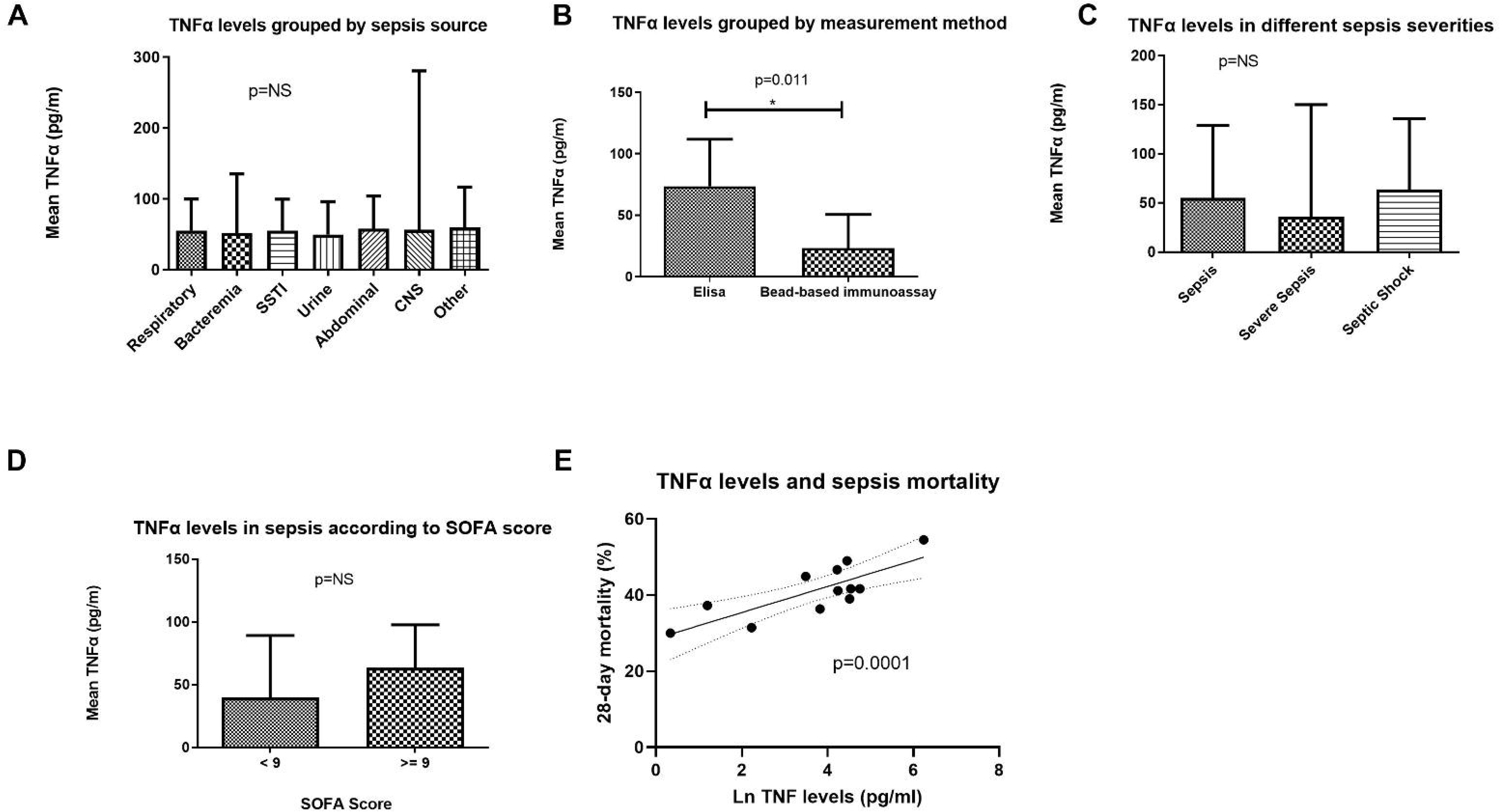
A: TNFα levels grouped by sepsis source. B: TNFα levels obtained using different quantification assays (p= 0.011). C: TNFα levels according to sepsis severity. D: TNFα levels stratified by Sequential Organ Failure Assessment (SOFA) SOFA score below 9 or greater than or equal to 9. E: TNFα levels and 28-day mortality. *p-value<0.05. SSTI= Skin and Soft Tissue Infections, CNS= Central Nervous System.

In a meta-regression, lower mean TNFα was observed in studies containing an increased percent of female patients and in studies containing sepsis patients with older age. Metaregression examining SOFA score showed SOFA did not associated with TNFα levels (Coef: -0.02, CI -0.4-0.3, p-value=0.931) (Figure 3D).

### TNFα Levels in Patients with Different Sepsis Severities and in Patients with Different Microbial Causes of Sepsis

There was no significant difference in mean TNFα level in patients with sepsis (55 pg/ml; 95% CI 23.5-128.9 pg/ml), severe sepsis (35.7 pg/ml; 95% CI 8.5-150.2 pg/ml), or septic shock (63.7 pg/ml; 95% CI 29.8-135.9 pg/ml; p= 0.783) (Figure 3C). The scatter plots showed no linear relationship between TNFα levels and SOFA score or year of study publication (supplement. eFigure 2, and eFigure 3, respectively). There was no significant difference in mean TNFα comparing reported participants with a SOFA score ≥ 9 (63.4 pg/ml; 95% CI 41.1-97.7 pg/ml) and participants with a SOFA score < 9 (39.7 pg/ml; 95% CI 17.7-89.2 pg/ml; p= 0.318) (Figure 3D).

### TNFα Levels and Sepsis Mortality

Elevated TNFα levels associated with increased 28-day mortality (p=0.001) (Figure 3E). The estimated 28-day percentage mortality increased as TNFα levels increased (Figure 3E. Supplement. Bubble plot, eFigure 1).

### TNFα Sensitivity Analysis

In an analysis of the effect of outlier studies on TNFα levels, one study was omitted, and a repeat analysis was conducted using the remaining 68 studies. This was repeated until each study in the meta-analysis was tested by being excluded. This manipulation did not significantly change the pooled TNFα concentration level of the remaining studies. The recalculated pooled TNFα levels ranged between 53.4 and 61.8 pg/ml. Excluding the largest outlier, Prashant’s(2013) study (unusually high TNFα level) decreased the pooled TNFα level the most to a calculated concentration of 53.4 pg/ml. There was also no significant change in heterogeneity after each study exclusion.

Furthermore, TNFα level effect sizes (pg/ml) did not change after decreasing the variance to 0.25. The levels decreased to 32.4 pg/ml, assuming an I^2^ of 10%. A funnel plot for publication bias indicated missing studies in the bottom left- and right-hand sides of the plot, suggesting asymmetry (supplement eFigure 4). The Egger test showed an estimated slope, β1, is 1.26 with a standard error of 1.38 (P=0.363), indicating a lack of small-studies effect.

### IL-1β Levels in Sepsis

Twenty-four studies reporting IL-1β levels in sepsis patients were included. The pooled estimated mean IL-1β level in sepsis patients was 21.8 pg/ml (95% CI 12.6-37.8 pg/ml; I^2^ =99.8%) (Figure 4). Among the different study types, prospective studies reported the highest mean IL-1β level in sepsis participants (35 pg/ml; 95% CI 13.4-91.6 pg/ml), while the mean IL-1β level in cross-sectional studies was the lowest (8.5 pg/ml; 95% CI 8.1-8.8 pg/ml; p= 0.005). There was no mean IL-1β difference comparing studies using different measurement techniques (28.8 pg/ml for ELISA and 12.5 pg/ml for bead-based immunoassay studies; p= 0.107). We also found no significant mean IL-1β difference comparing adult sepsis patients (25.6 pg/ml) with pediatric patients (9.7 pg/ml, p= 0.165). There were no statistically significant differences in IL1-β levels based on sepsis severity (sepsis, severe sepsis, or septic shock), SOFA score, or microbial sepsis source. Metaregression revealed no overall mortality association with IL-1β levels (Coef: -0.01, CI -0.05-0.03, p-value=0.59) and no association with 28-day mortality (Coef: -0.08, CI: -0.26-0.1, p=0.39).

**Figure 4:**
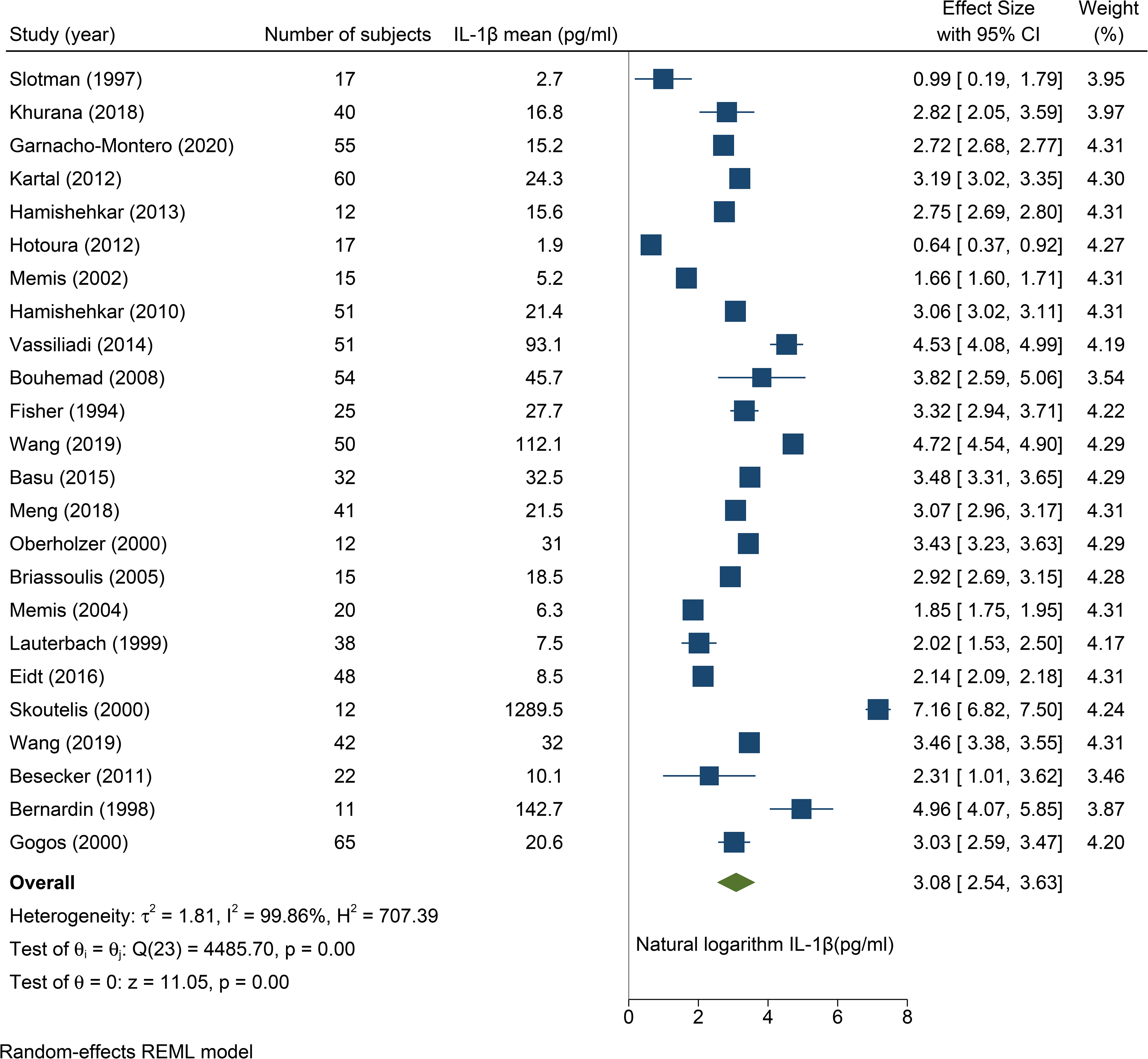
Forest plot for IL-1β studies [33, 39, 49, 57, 60, 62, 64, 71, 73, 80, 84, 87, 91, 93, 95–97, 100–105]. Effect size (pg/ml) is log-transformed

### IFNγ Levels in Sepsis

Five studies reporting IFNγ levels in sepsis patients satisfied inclusion criteria. The pooled estimate mean IFNγ level in sepsis patients was 63.3 pg/ml (95% CI 19.4-206.6 pg/ml; I^2^ = 99.7%) (supplement. eFigure 5). There were too few studies to perform subgroup or metaregression analysis for IFNγ.

### TNFα Levels in Healthy Person

Among studies in healthy persons, 12 disclosed TNFα levels in 611 participants. The pooled estimate mean TNFα concentration in health was 5.5 pg/ml (95% CI 3.8-8.0 pg/ml; I^2^ = 99.5%) (Supplement. eFigure 6). There were too few IL-1β studies or IFNγ studies to conduct meta-analyses.

## Discussion

In this systematic review and meta-analysis, the pooled mean TNFα level in sepsis was 58.4 pg/ml compared to 5.5 pg/ml in healthy volunteers. TNFα concentration correlated with the 28-day mortality in sepsis. Our review also found a mean IL-1β level of 21.8 pg/ml and a mean IFNγ concentration of 63.3 pg/ml in sepsis patients (Table 2). To our knowledge, this is the first systematic review and meta-analysis characterizing levels of key proinflammatory cytokines in the circulation in patients with sepsis.

**Table 2.** Summary of proinflammatory cytokine levels

|  | **Cytokine** | **Number of studies** | **Pooled estimate (mean pg/ml)** | **95% confidence interval (pg/ml)** |
| --- | --- | --- | --- | --- |
| **Sepsis patients** | TNFα | 69 | 58.4 | 39.8 – 85.8 |
| | IL-1β | 24 | 21.8 | 12.6 – 37.8 |
| | IFNγ | 5 | 63.3 | 19.4 – 206.6 |
| **Healthy persons** | TNFα | 12 | 5,5 | 3.8 – 8.0 |

Consistent with previously published studies, elevated TNFα levels are associated with increased mortality[20] [21]. Although we demonstrated association between increasing TNFα levels and 28-day mortality, numerous randomized placebo-controlled clinical trials failed to show a mortality benefit for TNFα-inhibiting therapies in patients with sepsis [12, 22, 23]. Most of these clinical trials did not quantify TNFα levels in sepsis patients. We believe two primary factors may account for the failure of anti-TNFα treatment. First, insufficient TNFα blockade is possible. However, this seems unlikely given the relatively low mean TNFα level obtained in sepsis patients in the present study. The mean TNFα level we determined does not appear to be excessive in magnitude and lowers the likelihood TNFα blocking power has been inadequate in TNFα-inhibition sepsis trials. Second, elevated TNFα could represent an epiphenomenon or non-causal associate of sepsis outcomes. In this case, TNFα elevations associate with disease severity but do not participate in harmful effects. We believe the latter is a more plausible explanation for the failure of TNFα-blocking agents to reduce sepsis mortality. If elevated TNFα levels caused mortality, TNFα likely exerts detrimental effects by causing end-organ damage. If so, we would expect higher TNFα to associate with elevated SOFA scores, a surrogate indicator of organ damage. However, we found no association between SOFA scores and TNFα levels.

There are large interstudy variations in sepsis patients’ proinflammatory cytokine levels, partly due to the measurement assay technique[24]. To mitigate this limitation, minimize measurement bias, and more closely reflect *in vivo* cytokine levels in the circulation, we restricted our inclusion criteria to studies using ELISA or a bead-based immunoassay. Enzyme-linked immunosorbent assay is likely the best-validated measurement technique for cytokines[25]. In a detailed study conducted by the World Health Organization, ELISA assays outperformed radioimmunoassay (RIA) in measuring TNFα in human serum, whereas RIA tended to overestimate TNFα levels[15]. Compared to ELISA, bead-based immunoassays have shown similar sensitivity for cytokine quantification[16, 26]. An additional consideration motivated the restriction of our analysis to these two assay methods. It seems likely future clinical studies will employ these assay platforms. This enhances the relevance of our results to clinical investigations moving forward.

The decades-long failure of cytokine antagonism to confer mortality benefit in sepsis suggests re-assessment of the concepts and assumptions underlying our approach to sepsis research, clinical investigation, and treatment. We believe there are two areas in critical need of examination. First, an updated and clinically-applicable definition of inflammation is needed. The absence of an empirically consistent understanding (or definition) of inflammation has led to a lack of focus in this field. For example, which cytokines are relevant for causing inflammation and tissue damage that characterize cytokine storm? With over 100 genes likely to code for cytokines[10], we need criteria that characterize which ones are relevant as causes of inflammation. Such a definition of inflammation should also differentiate inflammation from immune responses and unify clinical, histopathological, and molecular descriptions of inflammation. The second area in need of study is a comparative analysis of proinflammatory cytokines in sepsis and in non-sepsis diseases to gain much-needed perspective on the notion of an overabundant cytokine storm. Our results will permit us to answer a fundamental question regarding cytokine amounts in sepsis: “is that a lot?”

TNFα levels in seropositive rheumatoid arthritis can range between 47 pg/ml and 2,720 pg/ml [27, 28], and TNFα concentrations of 40 pg/ml have been measured in systemic lupus erythematosus [29]. TNFα has reached 862 pg/ml and 215 pg/ml in severe acute pancreatitis and salicylate-induced pseudosepsis, respectively [30, 31]. These markedly elevated levels suggest that TNFα levels in sepsis are not achieving “storming” levels as commonly thought. Moreover, it seems the term cytokine storm may be a misleading description of the cytokine response in any natural disease. Indeed, the belief that hypercytokinemia can directly induce lethality or organ malfunction seems dubious considering our data. Finally, this report is timely given the emphasis placed on cytokine storm in coronavirus disease 2019 (COVID-19) pathogenesis. Since severe COVID-19 is a form of sepsis, we believe re-assessment of the role of proinflammatory cytokine in COVID-19 pathogenesis is urgent.

## Limitations

This study has several limitations. There was significant heterogeneity in the data between studies explained by different study designs, measurement techniques, the timing of cytokine level measurement, and the characteristics of included patients. Some subgroup analyses were done based on the proportion of outcomes presented in each study, limiting confidence in derived associations. Missing data in some studies also limited the ability to reach more substantial statistical power. A possible publication bias exists due to the restriction of the included studies to English and the exclusion of unpublished studies. A small sample size of some studies reduces power to provide secure estimates of cytokine levels, especially for IL-1β and IFNγ. Finally, studies usually report cytokine levels at a single time point during the course of sepsis. Future studies should analyze the time-course of proinflammatory cytokines in sepsis using modern assay methods. This will reveal notable fluctuations in cytokine levels that may alter the interpretation of our results.

## Conclusions

In sepsis, circulating levels of TNFα are elevated approximately tenfold compared to levels in healthy persons. Concentrations of TNFα were linked to sepsis-related mortality but not to the severity of sepsis, organ dysfunction, or the microbiological cause of sepsis. Although cytokine levels are elevated in sepsis compared to healthy subjects, they appear to be similar to or lower than levels in non-lethal chronic inflammatory conditions. We need to revisit the paradigm of cytokine storm as the core mechanism of sepsis pathogenesis. We encourage follow-up studies that critically re-assess the causal role of elevated proinflammatory cytokines in sepsis.

## Data Availability

All data produced in the present work are contained in the manuscript.

## Author Contributions

Dr. Henao-Martínez had full access to all of the data in the study and took responsibility for the data’s integrity and the accuracy of the data analysis.

Concept and design: Shapiro, DeSanto, Franco-Paredes, Henao-Martínez.

Acquisition, analysis, or interpretation of data: Gharamti, Sillau, Samara, Monzon, DeSanto, Henao-Martínez, Shapiro

Drafting of the manuscript: Gharamti, Henao-Martínez, Shapiro

Critical revision of the manuscript for important intellectual content: Montalbano, Scherger, DeSanto, Chastain, Sillau, Montoya, Franco-Paredes, Henao-Martínez, Shapiro

Statistical analysis: Sillau, Henao-Martínez.

Administrative, technical, or material support: DeSanto, Henao-Martínez, Shapiro. Supervision: Franco-Paredes, Henao-Martínez, Shapiro.

## Supplementary Online Content

eMethods. Database search strategy

eFigure 1. Bubble plot TNFα levels and 28-day mortality

eFigure 2. TNFα levels and SOFA score scatter plot

eFigure 3. TNFα levels and year of study publication scatter plot

eFigure 4. TNFα levels publication bias funnel plot

eFigure 5. Forest Plot of studies with IFNγ levels

eFigure 6. Forest Plot of studies with TNFα levels in healthy persons

