## Supplementary material for "Proinflammatory Cytokine Levels in Sepsis and in Health and TNFα Association with Sepsis Mortality and patient characteristics: a Systematic Review and Meta-analysis"

**Supplementary Online Content**

eMethods. Database search strategy

eFigure 1. Bubble plot TNFα levels and 28-day mortality

eFigure 2. TNFα levels and SOFA score scatter plot

eFigure 3. TNFα levels and year of study publication scatter plot

eFigure 4. TNFα levels publication bias funnel plot

eFigure 5. Forest Plot of studies with IFNγ levels

eFigure 6. Forest Plot of studies with TNFα levels in healthy persons

**eMethods. Search strategy**

| MEDLINE (via Ovid MEDLINE^®^ ALL, 1946 to present) | |
| --- | --- |
| Search date = 1 May 2020 | |
| 1 | Sepsis/ or Shock, Septic/ or Purpura Fulminans/ or (sepsis or septic or purpura fulminans).tw,kf. |
| 2 | Interferon-gamma/ or (((interferon or IFN) adj1 (gamma or y)) or IFNgamma or IFNy).tw,kf. |
| 3 | Tumor Necrosis Factor-alpha/ or (((tumor necrosis factor or TNF) adj1 (alpha or a)) or TNFalpha or TNFα or ((tumor necrosis factor or TNF) adj1 soluble adj1 receptor adj2 ("1" or I or "2" or II)) or TNFsRp55 or TNFsRp75).tw,kf. |
| 4 | Interleukin-1beta/ or (((interleukin or IL) adj1 (1beta or 1b or 1-beta or 1-b)) or IL1beta or IL1β).tw,kf. |
| 5 | or/2-4 |
| 6 | Case-Control Studies/ or exp Clinical Trial/ or Clinical Trials as Topic/ or Controlled Clinical Trial/ or Cohort Studies/ or Cross-Sectional Studies/ or Longitudinal Studies/ or Prospective Studies/ or Randomized Controlled Trial/ or (case series or case-control or clinical trial or cohort study or cross-sectional or longitudinal study or prospective cohort* or (random* adj1 (clinical or control*) adj1 trial)).tw,kf. or randomized.ab. or placebo.ab. or randomly.ab. or trial.ti. |
| 7 | 1 and 5 and 6 |
| 8 | limit 7 to english language |
| 9 | exp animals/ not exp humans/ |
| 10 | 8 not 9 |
| 11 | limit 10 to yr="1985 -Current" |
| 12 | Healthy Volunteers/ or ((healthy or normal) adj1 (volunteer* or participant* or subject*)).tw,kf. |
| 13 | 11 and 12 |

**eFigure 1. Bubble plot TNFα levels and 28 days mortality**

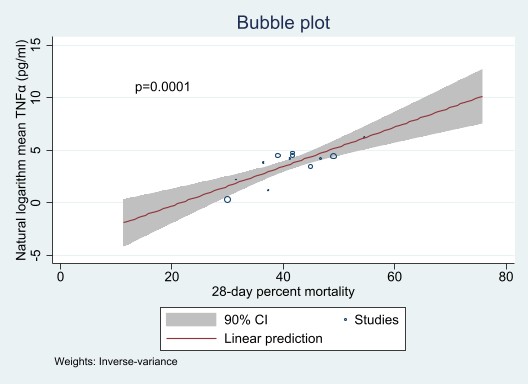

**eFigure 2. TNFα levels and SOFA score scatter plot**

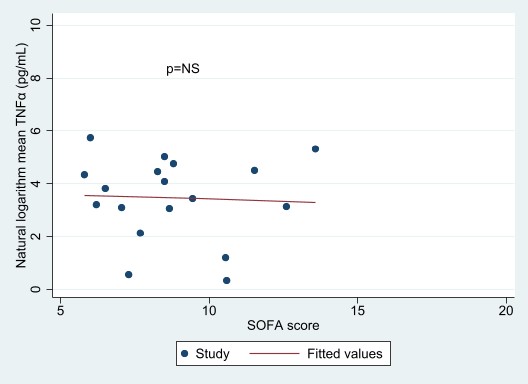

**eFigure 3. TNFα levels and year of study publication scatter plot**

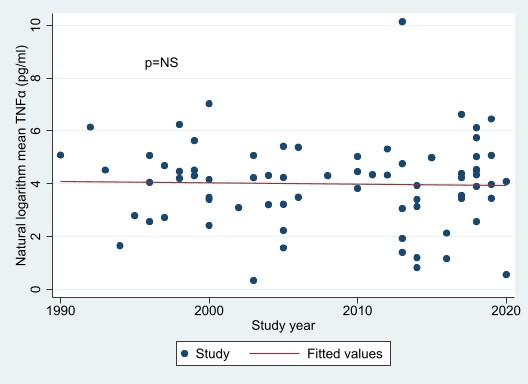

**eFigure 4. TNFα sepsis studies funnel plot**

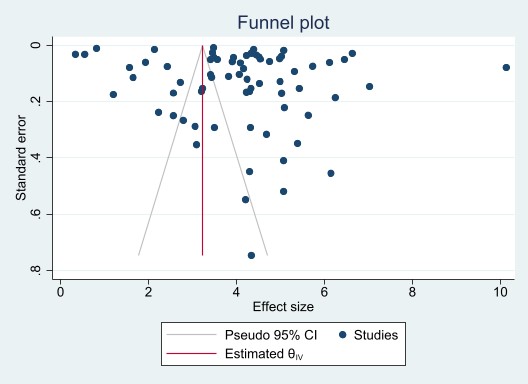

**eFigure 5. Forest Plot of sepsis studies with IFNγ levels**

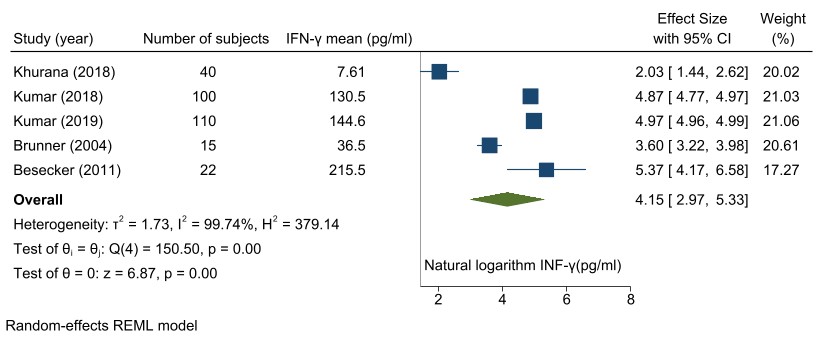

**eFigure 6. Forest Plot of sepsis studies with TNFα levels in healthy persons**

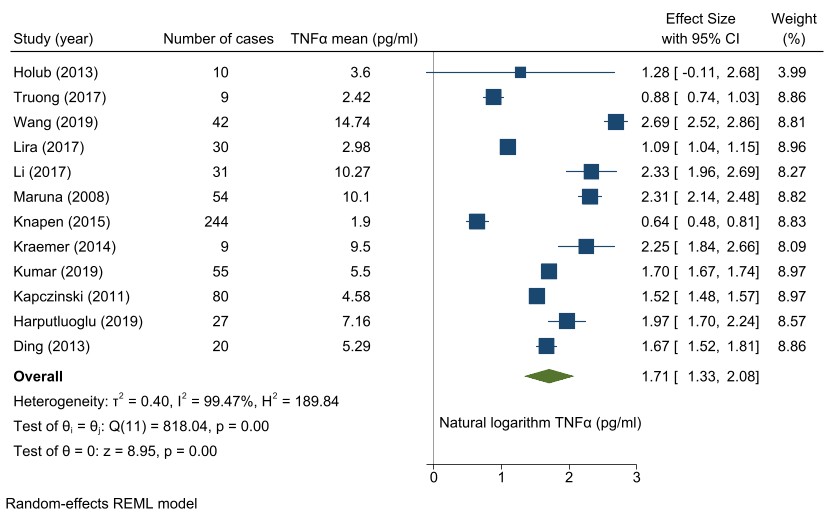
